# The International Classification of Functioning, Disability and Health to map leprosy-related disability in rural and remote areas in Indonesia

**DOI:** 10.1101/2023.07.21.23292986

**Authors:** Luh Karunia Wahyuni, Nelfidayani, Melinda Harini, Fitri Anestherita, Rizky Kusuma Wardhani, Sri Linuwih Menaldi, Yunia Irawati, Tri Rahayu, Gitalisa Andayani, Hisar Daniel, Intan Savitri, Petrus Kanisius Yogi Hariyanto, Isabela Andhika Paramita

## Abstract

The International Classification of Function, Disability and Health (ICF - WHO, 2001) recognizes several dimensions of disability, such as body structure and function (and impairment thereof), activity (and activity restrictions) and participation (and participation restriction) and their interactions with contextual factor (personal and environmental). In this study, we map and analyse the relationship between the components of ICF in leprosy patients from two rural areas in Indonesia: Lewoleba (East Nusa Tenggara) and Likupang (North Minahasa). This study was a part of a community outreach program by the KATAMATAKU team from Universitas Indonesia. The body structure was graded using the WHO hand and feet disability grade and the number of enlarged nerves, whilst the body function was measured by the Jebsen Taylor Hand Function Test (JTT) and Timed-up and Go (TUG). Activity limitation and participation restriction were measured using the Screening Activity Limitation Safety Awareness (SALSA) Scale and Participation Scale (P-scale), respectively. There were 177 leprosy patients from the two regions and 150 patients with complete data were included in the analysis. We found 82% subjects with multibacillary leprosy, 10.67% subjects with grade 2 WHO hand disability, and 9.33% subjects with grade 2 WHO foot disability. Assessment using SALSA Scale showed 29.33% of subjects with limitation activity and 11.33% with participation restriction. Age was shown to have positive correlations with SALSA, JTT, and TUG. Inter-dimensional analysis showed that the SALSA scale had significant positive correlations with the number of nerve enlargement, P-scale, JTT, and TUG. SALSA scores of grade 2 WHO hand and foot disability were also significantly higher than grade 1 and 0. Participation scale also had a positive correlation with JTT but not TUG. Hand disability seemed to affect societal participation whilst foot did not. We used the ICF to describe and analyse dimensions of leprosy-related disability in Indonesia.

**Author Summary:** Disability is the long-term outcome of untreated leprosy or Hansen’s disease, which is caused by peripheral nerve invasion of the *Mycobacterium leprae*. It is a serious and life-limiting complication to leprosy patients. Currently, there are seven regions in Indonesia which have not yet achieved the state of disease elimination. In addition, Indonesia has not succeeded in achieving the sub-target of rate of new cases with grade 2 disability in 2023. The concept of disability and health including the body structure, body function, personal and environmental domains, activity limitation, and participation restriction, is provided by the ICF. Through the framework, we could assess the aspects of disability and its relationship to one another in leprosy patients. Furthermore, the early detection, screening, and management programs for leprosy have not included aspects of disability and rehabilitative measures for restoration. Thus, the purpose of this study is to find the impact of leprosy-related disability in the patients’ body function, activity, and participation. We report 150 leprosy patients from two rural areas of Indonesia and found significant relationships between specific indicators, which would be useful for rehabilitative management programs in the future.

## Introduction

Infiltration of *Mycobacterium leprae* into Schwann cells in leprosy can cause peripheral nerve inflammation and subsequent progressive loss of nerve fibre function or neuropathy.[1] This condition mainly affects the hands (44.45%), feet (39,76%), and face (15,74%).[2] The subsequent loss of sensory; motor; and autonomic nerve function would appear as loss of thermal, nociceptive, and pressure senses; muscle paresis; and dryness of the skin, respectively. Occasionally, delayed emergence of skin lesions and sensory impairment may also occur, termed as silent neuropathy.[1] When left untreated, these conditions can develop into visible physical deformities (lagophthalmos, severe visual impairments, ulcers, clawing and shortening of digits in the extremities), which the World Health Organization (WHO) classify as grade 2 disabilities.[3] Unfortunately, the majority of newly diagnosed leprosy patients have already had deformities.[2]

Disabilities in leprosy patients cause physical disability, psychological disturbances, and extensive loss of manpower and economic loss to the society.[3] Assessment of disability in leprosy patients is a very important factor in the evaluation of the effectiveness of a leprosy elimination program.[3] Disability can be perceived as any impairment of a person’s health and mind condition that render the person to have functioning difficulty at the body, person, or societal levels, in any life domain.[4,5] The International Classification of Functioning, Disability and Health (ICF) recognizes dimensions of disability, which include body structure and function (along with each’s impairments), activity and limitations, as well as participation and restrictions. Environmental aspects of the individual are also emphasized as a major predictor of disability outcomes thereby shifting the paradigm of disability as a mere ‘medical’ or ‘biological’ dysfunction.[4]

Along with India and Brazil, Indonesia is one of the major leprosy contributors in the world. In the 2021 global leprosy update by WHO, the majority of new case detection came from South-East Asia and more than 10,000 cases were found in Indonesia.[6] According to the Indonesian Ministry of Health report, leprosy cases are still increasing annually, from 5 cases per 100.000 population in 2021 to 5.5 cases per 100.000 population in 2022.[7] Recent reports have shown that the number of leprosy patients with grade 2 WHO disability has widely ranged from 6.31% to 68.2%.[2,8,9] As of 2021, there were 12,230 registered leprosy cases in Indonesia, with a new case detection rate of 4.03 per 100,000 population and 10,976 new cases reported. The proportion of new leprosy cases with grade 2 deformities was determined to be 2.46 per 1,000,000 population, which still surpasses the WHO’s target of 0.92 per 1,000,000 population by 2023.[10,11] However, studies related to the burden of leprosy are fewer than those related to endemicity.[12] Not to mention that research pertaining to the specific challenges encountered by leprosy-related disabilities patients in Indonesia are very scarce. These data are urgently needed for initiating sustainable rehabilitative measures for disability prevention and for improving the patients’ well-being.

In this occasion, we want to present a study to analyze factors affecting leprosy-related disabilities involving participants from two remote areas in Indonesia: Lewoleba, Lembata, East Nusa Tenggara and Likupang, North Minahasa, North Sulawesi.

Lewoleba, which is the capital of Lembata island, East Nusa Tenggara, is in the Nubatukan district. Lewoleba has an area of 165.64 KM^2^ whilst Lembata spans over 1.266,4 KM^2^. Lembata island is divided into nine districts and has overall a tropical climate with a long drought. Lembata has relatively high humidity at around 80%.[13] Nubatukan district has the highest population density at 50.984 or 37% of total population of the island. Around 11.4% of total population are aged 0-4 years. There are three hospitals and one public health center in Nubatukan.[14] The population in Lembata mostly work in agriculture, forestry, hunting and fisheries sectors.[14] East Nusa Tenggara is considered as one of the provinces in Indonesia with high leprosy incident, with around 7 per 100.000 people and high chance of getting disability due to leprosy about 27.78%.[14,15]

Likupang region is further divided into three subdistricts: East Likupang, West Likupang and South Likupang. All of them are coastal subdistricts located at 1.6720° N, 125.0553° E, 0 meters above the sea level, with a total area of 298.27 square kilometer (KM^2)^.[16,17,18] Since Likupang is located near the seashore, it has high humidity, reaching 80-90% on average.[19] In 2022, the region had 44.660 inhabitants, with average population growth rate of 1.39% between the year 2020 and 2022. South Likupang is the subdistrict with highest population density, while East Likupang is the lowest.[20] According to data from Ministry of Health in 2021, North Sulawesi is one of Provinces in Indonesia with high leprosy cases, with detection rate of 11 per 100,000 inhabitants in 2018.[21,22] As of 2022, North Sulawesi is one of the 7 provinces where leprosy has not yet been eliminated. Likupang itself has 4 public health centers, 3 with inpatient care, as well as 3 clinics spread over three subdistricts. [16,17,18]

The objectives of this study are to report leprosy cases from rural and remote areas in Indonesia, map the functional disabilities of individuals diagnosed with leprosy in Likupang and Lewoleba, Indonesia, using ICF, and to find correlation between each aspect of ICF in Indonesian population.

## Materials and methods

### Ethical aspects

The study protocol was approved by the Ethics Committee of Faculty of Medicine, Universitas Indonesia under ethical clearance ND-454/UN2.F1/ETIK/PPM.00.02/2022 and was designed in consideration of the principles proposed by the Helsinki Declaration. Written formal consent was obtained directly from participants.

### Study design and population

This cross-sectional study was a part of a community outreach program by KATAMATAKU and was carried out in Lewoleba, Lembata, East Nusa Tenggara (July 2022) and Likupang, North Minahasa, North Sulawesi (August 2022). The KATAMATAKU team is a multidisciplinary (expert) team made up of dermatologists, ophthalmologists and physical medicine and rehabilitation (PMR) specialists from Faculty of Medicine Universitas Indonesia in Jakarta, Indonesia. The word KATAMATAKU is an Indonesian acronym for ‘*Identifi<u>ka</u>si <u>ta</u>nda-tanda <u>ma</u>ta, ekstremi<u>ta</u>s dan <u>ku</u>lit pada kus<u>ta</u>’* or identification of ocular, extremities, and dermatological signs in leprosy. The collaborative team often visits remote sites with higher leprosy-population density to do surveillance and charitable community engagements with leprosy patients and survivors. The outreach program consisted of physical examination, consultation, and treatment for leprosy patients. We contacted the local public health officers a month prior to the event and the affiliated regional hospitals and primary health care facilities notified the residing population nearby and registered leprosy patients.

The study participants from Likupang, North Minahasa and Lewoleba, Lembata were those who attended the events held in July 2022 and August 2022, respectively. Patients with leprosy who had provided written consent to take part in this research were eligible for inclusion. Each participant filled out the SALSA questionnaire as well as the participation scale (P-scale) and went through the entire assessment process, including the timed up and go (TUG) test and the Jebsen-Taylor Hand Function Test (JTT).

### Study protocol

In both Likupang and Lewoleba, information was gathered through the examination of all leprosy patients. All subjects who agreed to participate in the study were given face-to-face interviews with close-ended question forms. Age, gender, and educational background were among the basic demographic data collected. The study variables or assessment tools along with each’s descriptions are listed in **Table 1**. The outcomes were analysed in accordance with the International Classification of Functioning, Disability and Health (ICF) domains as listed in **Figure 2**.

**Figure 1.**
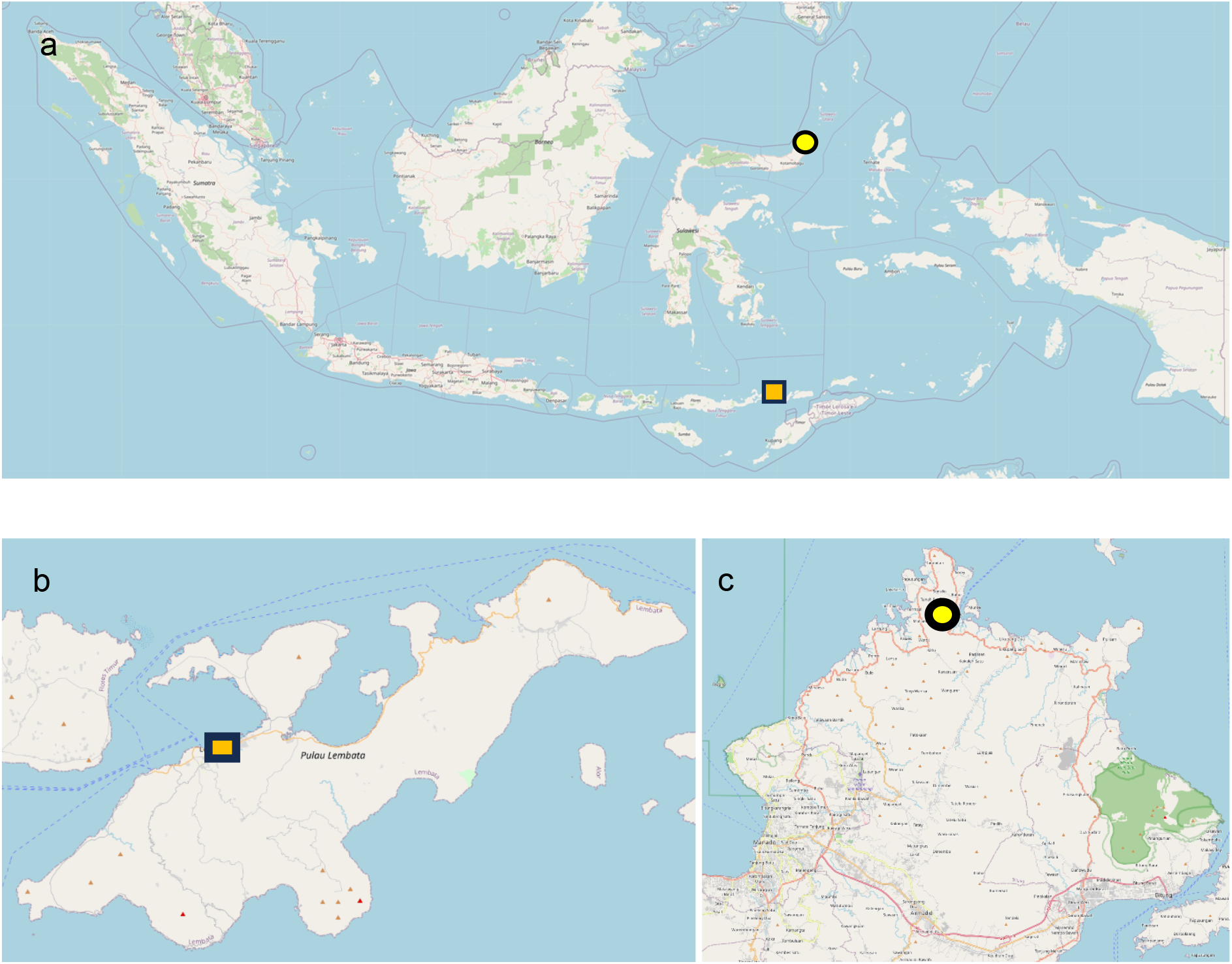
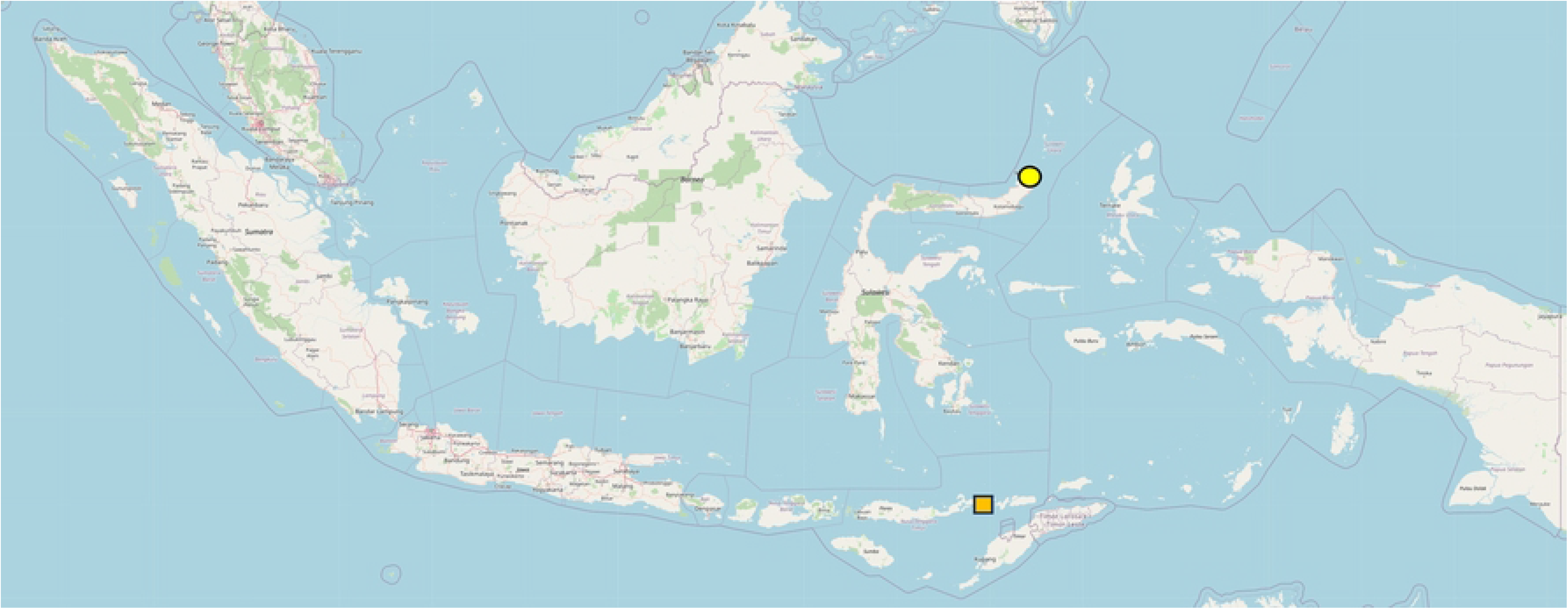
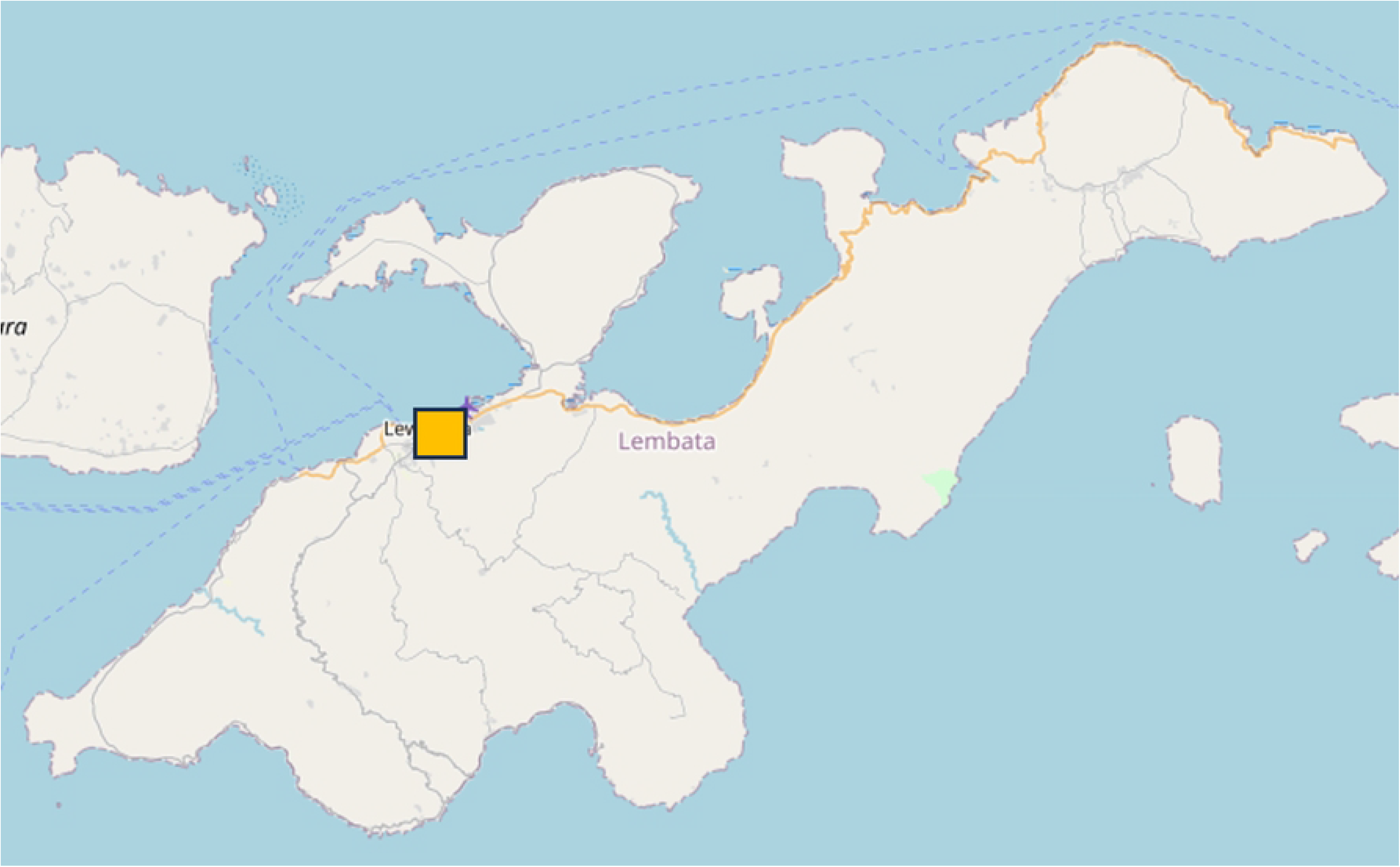
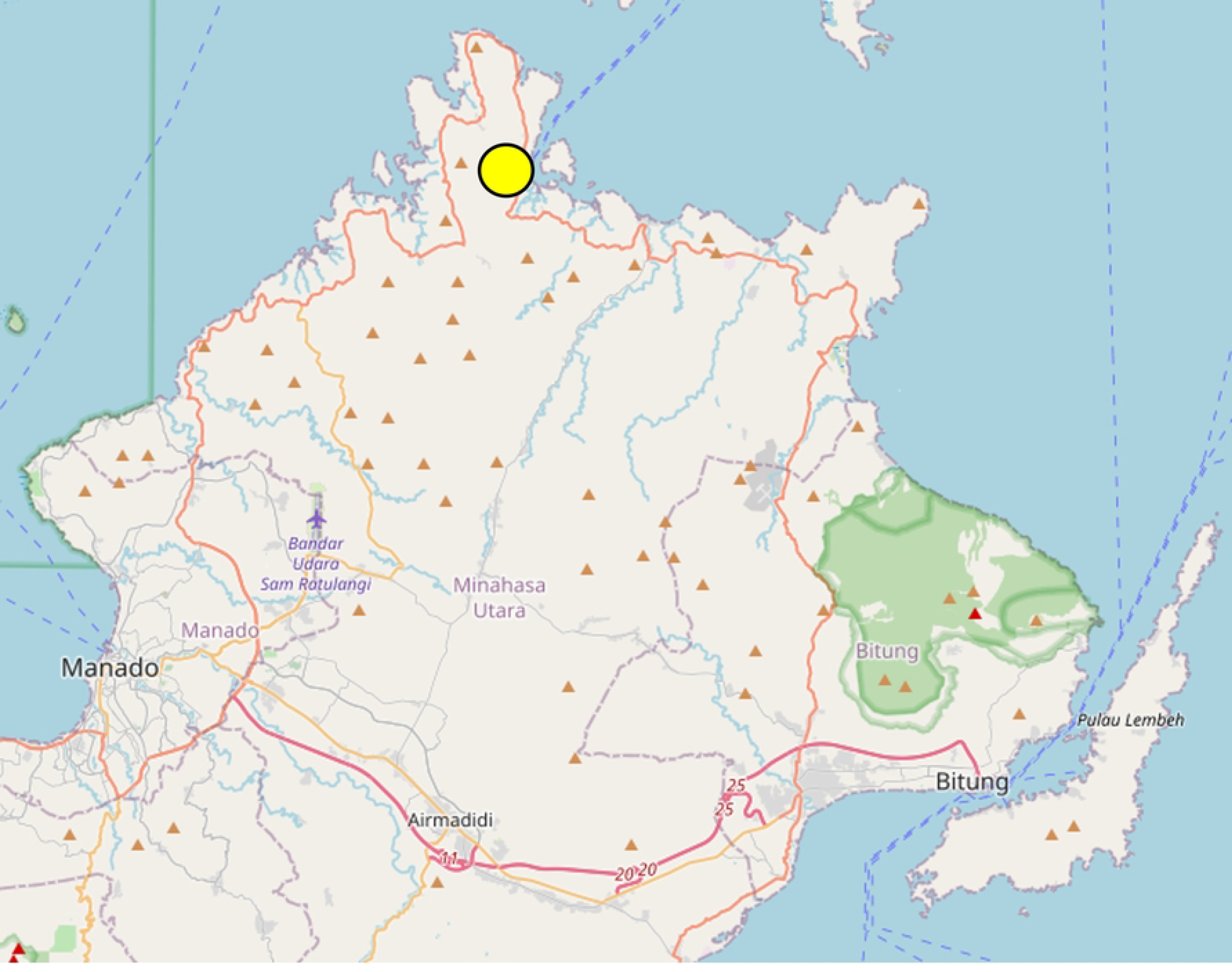
Map of Lewoleba (Lembata, East Nusa Tenggara) and Likupang (North Minahasa, North Sulawesi), Indonesia. (A) Map of Indonesia, scaled 1: 5900000 (1062×460), yellow circle indicates Likupang, North Minahasa and orange rectangle indicates Lewoleba, Lembata. (B) Map of Lembata Island (East Nusa Tenggara), scaled 1: 273897 (1077 x 658), orange rectangle indicates Lewoleba. (C) Map of North Minahasa (North Sulawesi), scaled 1: 138380 (1341 x 1077), yellow circle indicates Likupang. The map contains information from OpenStreetMap and OpenStreetMap Foundation, which is made available under the Open Database License (accessed online on 13^th^ of July 2023). Manual markings were done to locate Lewoleba and Likupang.

**Figure 2.**
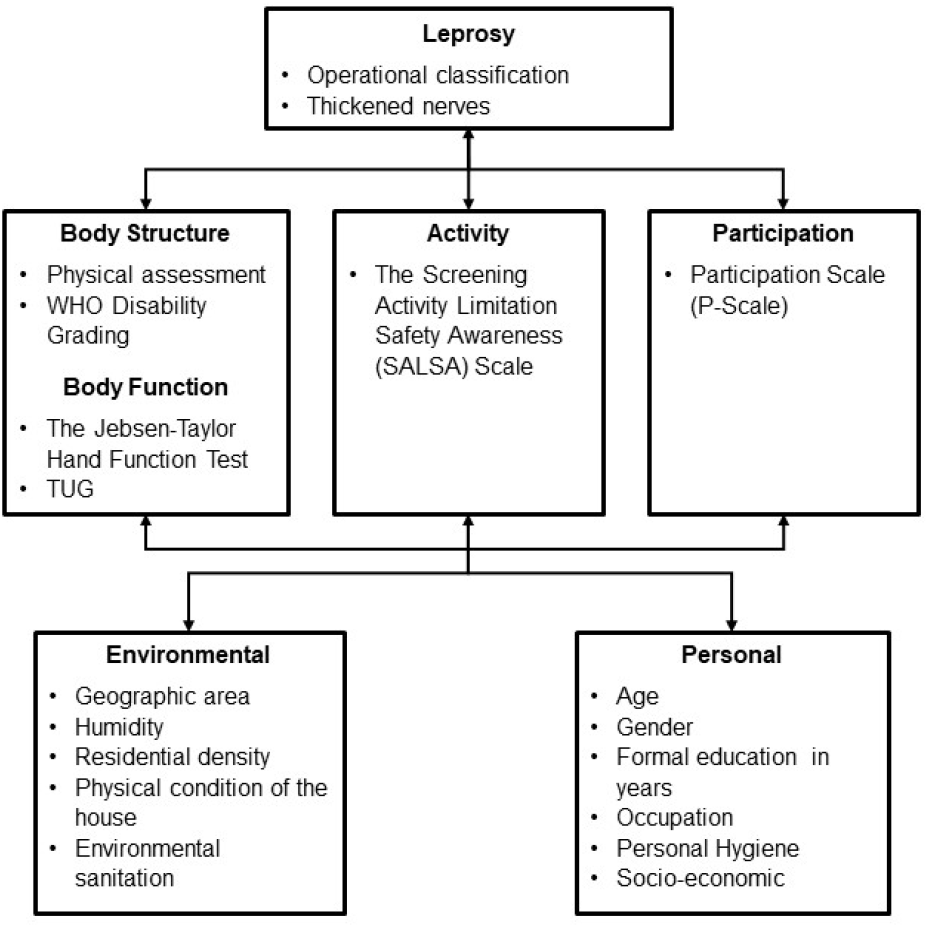
Application of the International Classification of Functioning, Disability and Health (ICF) domains in the study.[23].

**Table 1.** Assessment Tools.

| Item | Content | Evaluation | Note |
| --- | --- | --- | --- |
| *Personal factors* |  |  |  |
| Age |  | Identity card | <18 years old, 18-59 years old, ≥60 years old |
| Gender |  | Identity card | Female, male |
| Formal education level |  | Question to the patient | - Uneducated<br>- Primary Education<br>- Middle School<br>- Secondary Education<br>- Higher Education |
| *Health condition* |  |  |  |
| Diagnosis of Leprosy |  | Anamnesis related to history of the disease, type of leprosy, and medication | Paucibacillary, multibacillary |
| Number of person(s) living together with the same diagnosis |  | Anamnesis of any contacts within family members and whether they live together or separately with the patient | Number of family member with similar symptoms or diagnosis |
| *Body structure* |  |  |  |
| Physical Examinations | Abnormalities and deformities in the skin and extremities | - Inspection of lesions and deformities<br>- Motor and sensory nerve function | Number of nerve thickening. |
| The WHO classification of physical disability in leprosy.[24] |  | The presence of nerve impairments or visible deformity | Grade 0: no disability (no loss of sensitivity and no visible deformity or damage to the eyes, hands or feet) |
|  |  |  | Grade 1: only disability (a loss of sensitivity without visible deformity or damage to the eyes, hands or feet)<br>Grade 2: the presence of visible deformity or damage to the eyes (lagophthalmos, iridocyclitis, corneal opacities, severe visual impairment), hands (claw hands, ulcers, absorption of the digits, thumb-web contracture and swollen hand), feet (plantar ulcers, footdrop, inversion of the foot, clawing of the toes, absorption of the toes, collapsed foot and callosities) |
| *Body function* |  |  |  |
| The Jebsen-Taylor Hand Function Test (JTT).[25] | A standardized and objective measure of fine and gross motor hand function using simulated activities of daily living (ADL) | A series of seven subtests representing fine motor, non-weighted and weighted hand function in ADL, which includes: writing, simulated page-turning, lifting small objects, simulated feeding, stacking, and lifting large, lightweight, and heavy objects. | The subtests are scored by recording the number of seconds required to complete each test<br>Total score is the sum of time taken for each sub-test, which are rounded to the nearest second. Shorter times indicate better performance. |
| Timed Up and Go Test (TUG).[26] | To determine the patient's fall risk and measure the progress of balance, sit to stand and walking | Assessed domains<br>• Balance<br>• Gait<br>• Walking speed | Patients performed the test one time, and if a clear error was made, they were asked to repeat the TUG. |
| *Activity* |  |  |  |
| The Screening Activity Limitation Safety Awareness (SALSA) Scale.[27,28] | Includes 20 daily activities questions related to the three areas of mobility, self-care, and work. | The ability of patients to do basic daily activities such as walking, self-care, carrying heavy objects and manipulating small objects. | SALSA's Indonesian edition was used for this investigation. The total score ranges from 0-80, with the result classified as:<br>• no substantial limitation (0-24)<br>• mild limitation (25-39)<br>• moderate limitation (40-49)<br>• severe limitation (50-59)<br>• extreme limitation (60-80) |
| *Participation* |  |  |  |
| Participation Scale (P-Scale).[29] | Consists of 18 items to measure (social) participation for use in rehabilitation, stigma reduction and social integration programmes. | Assesses domains of participation including:<br>• Communication<br>• Mobility<br>• Self-Care<br>• Domestic Life<br>• Interpersonal Interactions | The P-scale total score varies between 0-90, with the result classified as:<br>• No significant restriction (0-12)<br>• Mild restriction (13-22)<br>• Moderate restriction (23-32)<br>• Severe restriction (33-52) |
|  |  | • Major life areas (work, education, etc)<br>• Community, Social and Civic | • Extreme restriction (53-90) |

Microsoft Excel was used to store, organize, clean raw data and perform F tests for equal variance. Statistical analysis was performed using GraphPad Prism version 10.0.0. Normality and F tests were performed to all sets of data. For the comparison test between two groups, non-normal groups will be analysed non-parametrically with Kolmogorov-Smirnov test if both groups have equal variances. Groups with non-equal variances will be analysed using Mann-Whitney U test. For the comparison between more than two groups, groups with non-normal distribution will be analysed using Kruskal-Wallis test. Bivariate analyses using Pearson’s and Spearman’s correlation tests were performed in normally and non-normally distributed variables, respectively. Categorical variables are analysed using Fisher’s Exact and Chi-Square tests. Probability of 0.05 was considered statistically significant.

## Results

The total number of patients examined during the events were 177 patients with leprosy. After excluding patients with incomplete data, 150 patients (107 in Lewoleba and 43 in Likupang) were included in the analysis. Mean age of the patients was 44.71 ± 18.7 years. Majority of the patients were between 18-59 years old, male, graduated from elementary/primary schools, did not have hand or foot disability, have no limitation nor restriction in functional activity and participation, respectively, had been diagnosed with multibacillary type, and had none other family members living together diagnosed with leprosy (**Table 2**).

**Table 2.** Demographics of Leprosy Patients in Lewoleba (Lembata) and Likupang (North Minahasa), Indonesia (n=150).

| Characteristics | n | % |
| --- | --- | --- |
| *Age* |  |  |
| < 18 years | 13 | 8.67 |
| 18-59 years | 97 | 64.67 |
| > 60 years | 40 | 26.67 |
| *Gender* |  |  |
| Female | 46 | 30.67 |
| Male | 104 | 69.33 |
| *Educational Background* |  |  |
| Uneducated | 9 | 6 |
| Primary Education | 80 | 53.33 |
| Middle School | 21 | 14 |
| Secondary Education | 33 | 22 |
| Higher Education | 7 | 4.67 |
| *WHO Hand Disability* |  |  |
| Grade 0 | 114 | 76 |
| Grade 1 | 20 | 13.33 |
| Grade 2 | 16 | 10.67 |
| *WHO Foot Disability* |  |  |
| Grade 0 | 110 | 73.33 |
| Grade 1 | 26 | 17.33 |
| Grade 2 | 14 | 9.33 |
| *SALSA Scale* |  |  |
| No limitation | 106 | 70.67 |
| Mild limitation | 41 | 27.33 |
| Moderate limitation | 2 | 1.33 |
| Severe limitation | 0 | 0 |
| Extreme limitation | 1 | 0.67 |
| *Participation Scale* |  |  |
| No restriction | 133 | 88.67 |
| Mild restriction | 10 | 6.67 |
| Moderate restriction | 6 | 4 |
| Severe restriction | 0 | 0 |
| Extreme restriction | 1 | 0.67 |
| *Types of Leprosy* |  |  |
| Paucibacillary | 27 | 18 |
| Multibacillary | 123 | 82 |
| *Number of family members with leprosy living together* |  |  |
| None | 103 | 68.67 |
| 1 | 38 | 25.33 |
| 2 | 5 | 3.33 |
| > 2 | 3 | 2 |
| Undisclosed | 1 | 0.67 |

Analysis of the data were done according to ICF, introduced by the WHO in 2001. We divided the examinations according to several aspects. Age, gender, and types of leprosy are included into the patient’s condition. The number of thickened nerves and WHO hand and foot disability grading represent body structure. Jebsen-Taylor Hand Function Test and TUG are parts of body function. The SALSA and Participation Scales represent activity and participation, respectively. Initially, we would like to find whether there are any differences of patient’s condition, body structure, body function, activity, and participation according to gender, age, education, and types of leprosy. All data were not normally distributed, and many sets of groups did not have equal variances. We found a significant difference of the number of thickened nerves between male and female (Mann-Whitney U test, median 0 vs 0, p=0.0021), as the SALSA, P-scale, JTT, and TUG were not. There were significant positive correlations between age and SALSA scales including the mobility (r=0.3965, p<0.0001), work (r=0.2939, p=0.0003), and dexterity (r=0.4738, p<0.0001) aspects, JTT in both hands, and TUG as seen in **Figure 3A-C**. Finally, we also found that the SALSA scales in different educational level were also found to be significantly distinctive (Kruskal-Wallis test, p<0.0099), whereas participation scales were not (**Table 3**).

**Figure 3.**
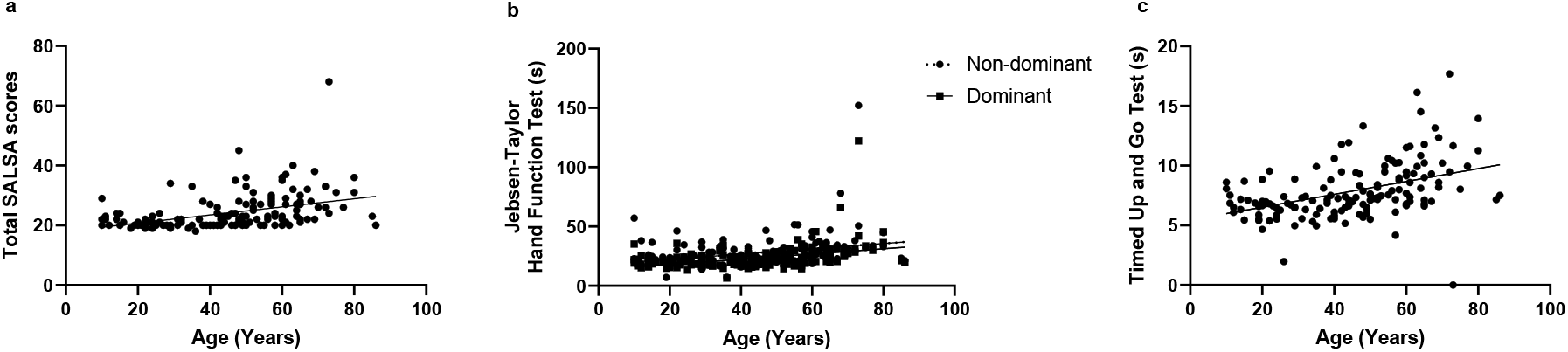
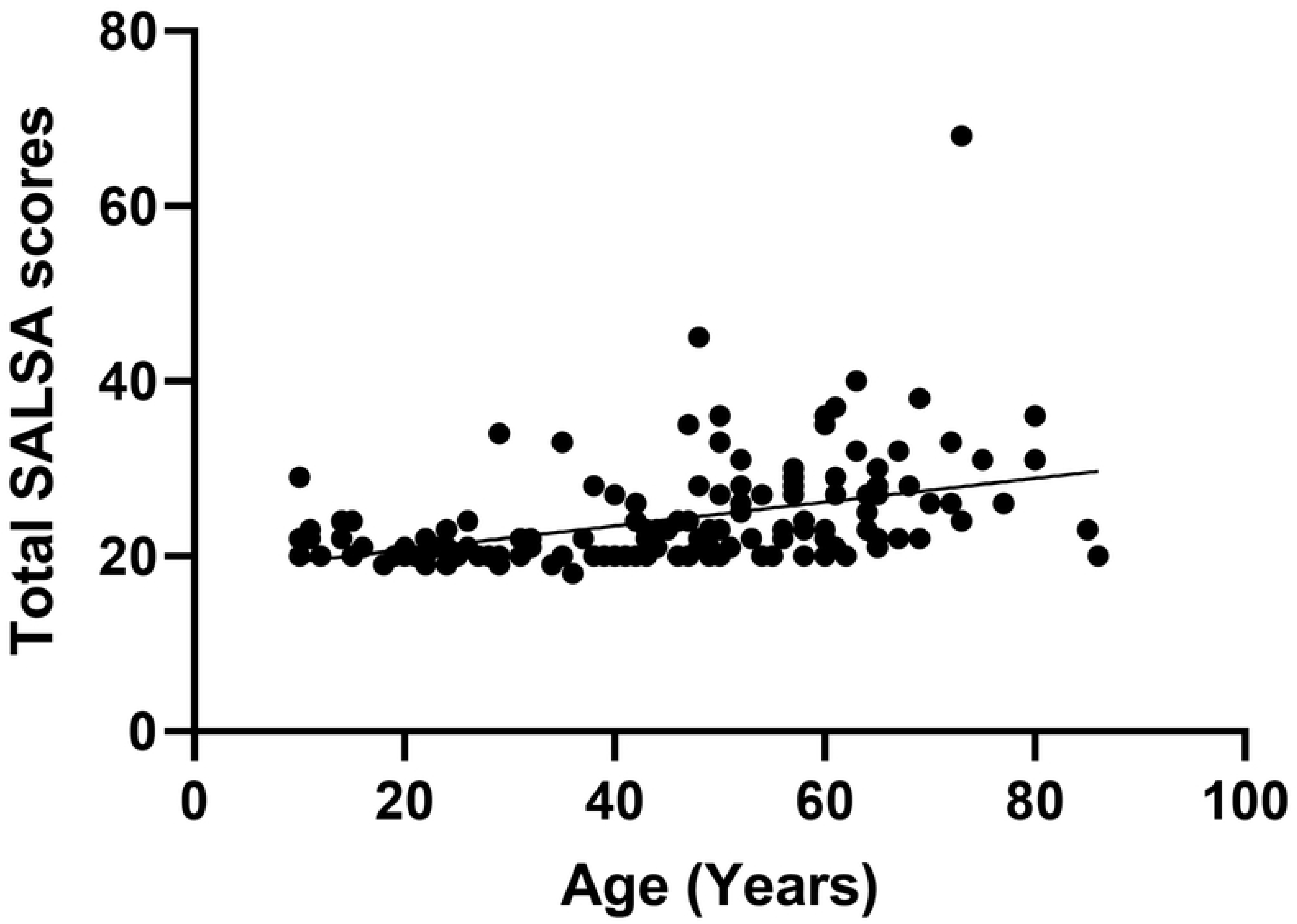
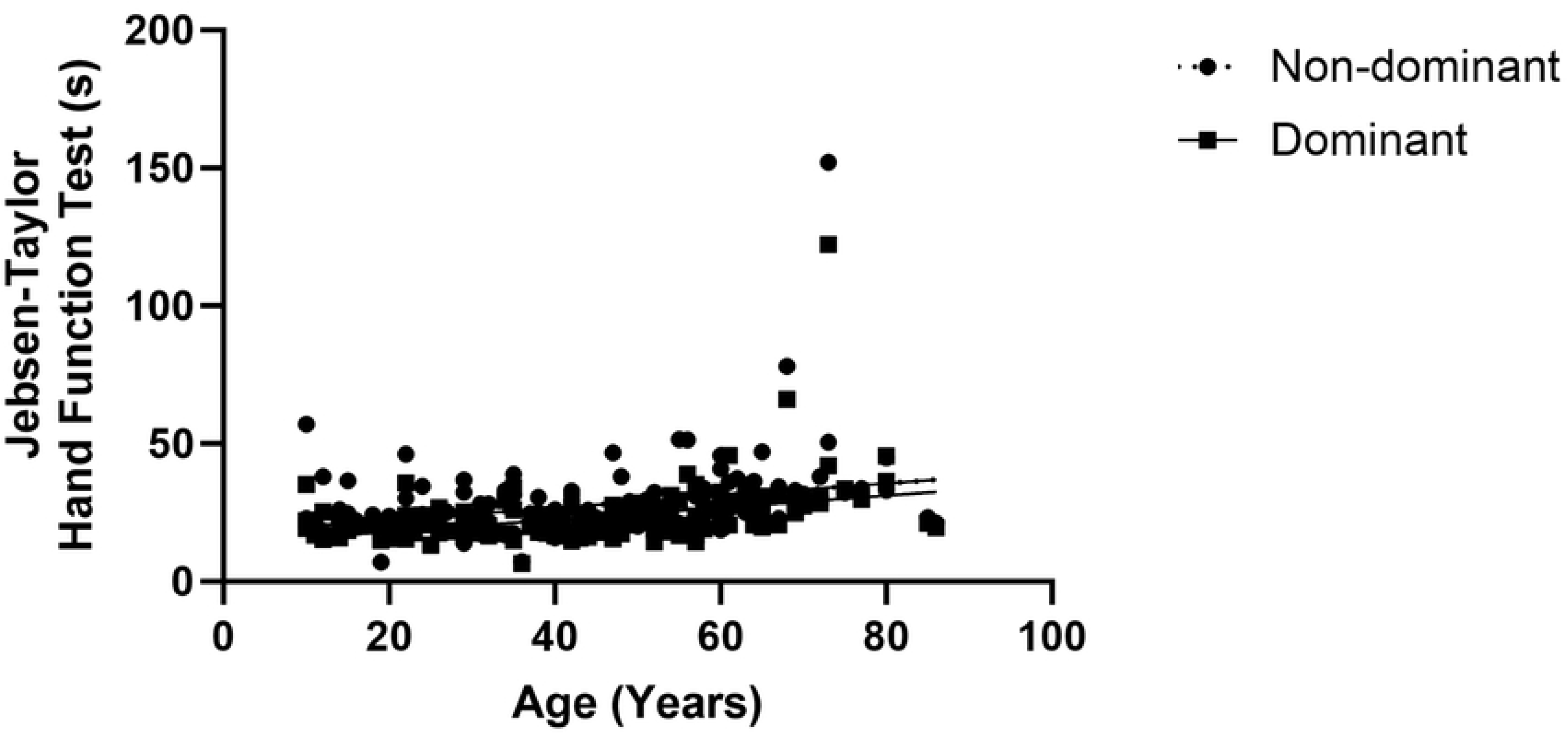
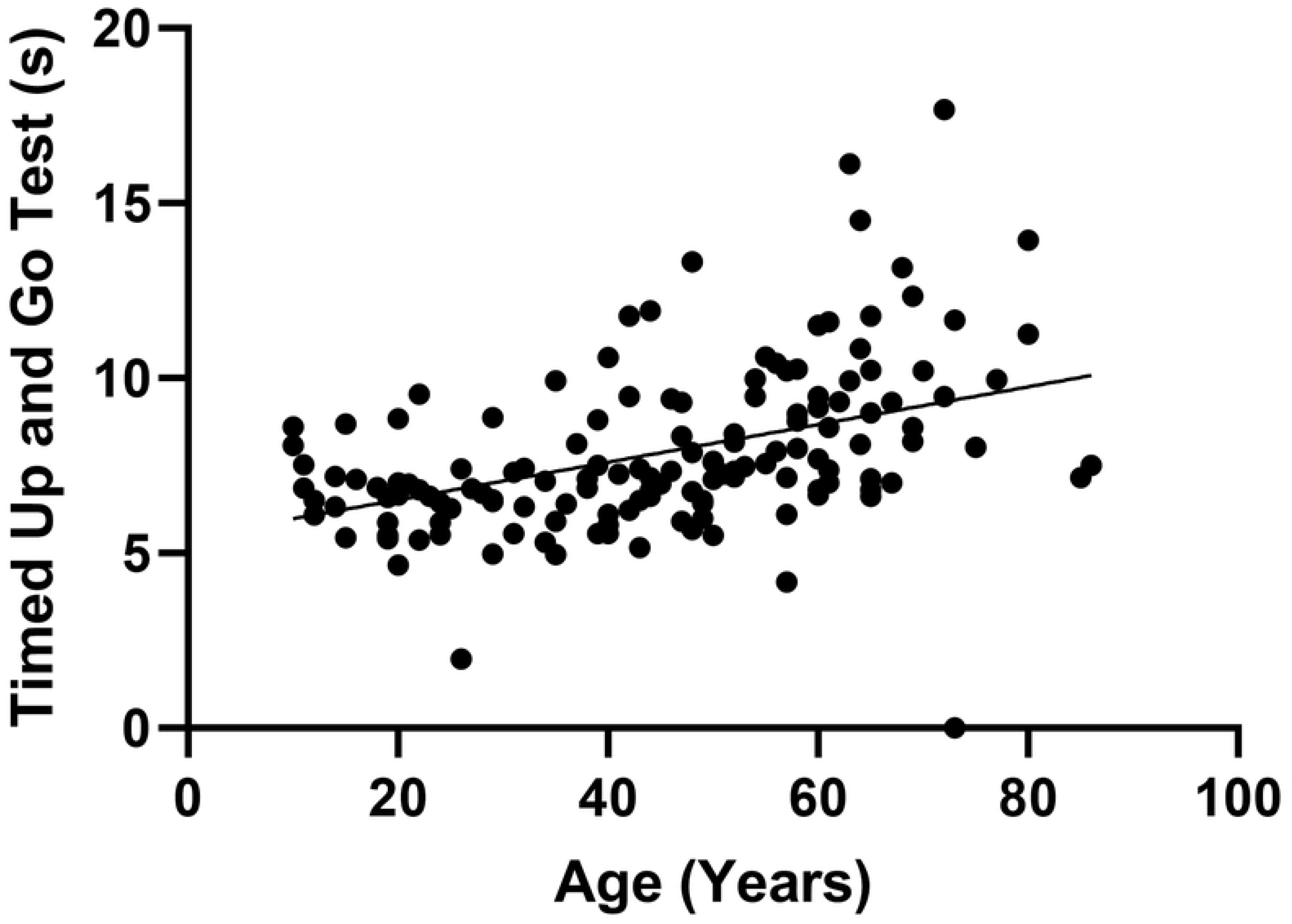
Significant bivariate analyses of age and activity or body function. X-axis is age defined in years, Y-axis is listed as follows; (A) total SALSA scores (Spearman’s r=0.5047, p<0.0001), (B) JTT in non-dominant and dominant hands (Spearman’s r=0.4203, p<0.0001 vs Spearman’s r=0.4942, p<0.0001, respectively), C: TUG (Spearman’s r=0.4904, p<0.0001). SALSA, screening of activity limitation and safety awareness; JTT, Jebsen-Taylor hand function test; TUG, timed up and go test.

**Table 3.** Comparison of SALSA and P-scales according to different educational level using Kruskal-Wallis test.

| Assessment Tools | Uneducated | Primary Education | Middle School | Secondary Education | Higher Education | p-value |
| --- | --- | --- | --- | --- | --- | --- |
| **SALSA (median)** | 22 | 23 | 21 | 21 | 22 | 0.0099* |
| **P-scale (median)** | 1 | 1 | 0 | 1 | 0 | 0.5738 |

\* Significant value

To assess different domains in relation to body structure impairment, Kruskal-Wallis tests were performed on SALSA scores, P-scales, JTT, and TUG according to WHO hand and foot disability grades. We found significant differences of the medians of SALSA scale, P-scale, and JTT in patients with grade 0, 1, and 2 WHO hand disability, whilst only comparison of SALSA scales in patients with grade 0, 1, and 2 WHO foot disability yields significant results as seen in **Table 4 and 5**.

**Table 4.** Comparison of hand structure impairment according to hand function impairment, activity limitation, and participation restriction using Kruskal-Wallis test.

| WHO Hand Disability | SALSA Scale (Median) | SALSA-Work (Median) | SALSA-Dexterity (Median) | P-scale (Median) | JTT Non-Dominant (s) (Median) | JTT Dominant (s) (Median) |
| --- | --- | --- | --- | --- | --- | --- |
| Grade 0 | 21.5 | 7 | 5 | 1 | 24.37 | 20.56 |
| Grade 1 | 23.5 | 7 | 6 | 3 | 26.24 | 25.02 |
| Grade 2 | 27.5 | 8.5 | 8 | 1.5 | 35.62 | 24.51 |
| p-value | 0.0012* | 0.0276* | 0.0065* | 0.0087* | 0.003* | 0.0137* |

\* Significant value

**Table 5.**
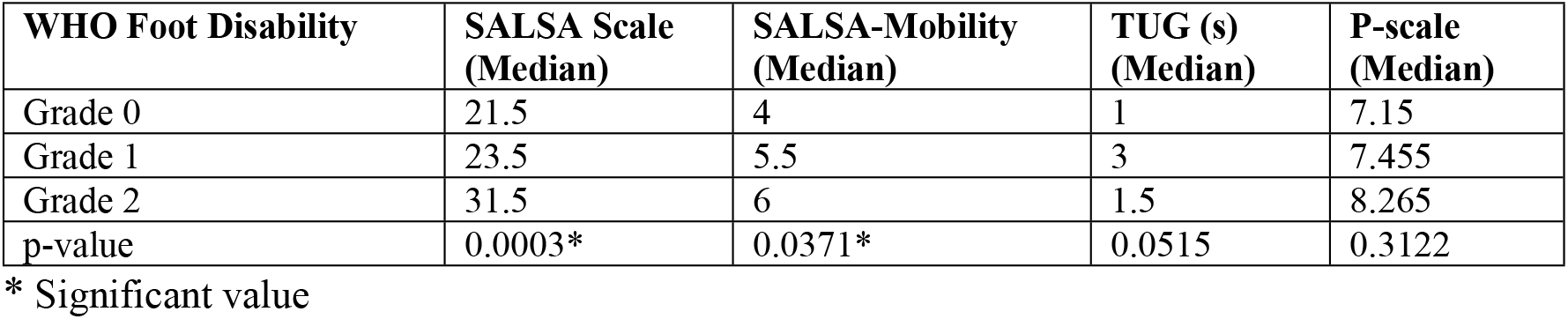
Comparison of foot structure impairment according to foot function impairment, activity limitation, and participation restriction using Kruskal-Wallis test.

| WHO Foot Disability | SALSA Scale (Median) | SALSA-Mobility (Median) | TUG (s) (Median) | P-scale (Median) |
| --- | --- | --- | --- | --- |
| Grade 0 | 21.5 | 4 | 1 | 7.15 |
| Grade 1 | 23.5 | 5.5 | 3 | 7.455 |
| Grade 2 | 31.5 | 6 | 1.5 | 8.265 |
| p-value | 0.0003* | 0.0371* | 0.0515 | 0.3122 |

\* Significant value

Finally, we would like to find correlation between each domain of ICF. Comparison tests of body structure impairment, body function impairment, activity limitation, and participation restriction between types of leprosy; paucibacillary or multibacillary, were performed. Variables with numerical data were compared non-parametrically using Kolmogorov-Smirnov and Mann-Whitney U tests and no significant differences were found. Associations between categorical variables were calculated using Fisher’s Exact and Chi-Square tests with no statistically significant results. We then performed statistical tests between other variables and found weak but significant positive correlation between total SALSA scores and the number of nerve enlargement (**Figure 4A**). There were also positive and significant correlations between P-scale and the number of thickened nerves, SALSA scale, which includes the SALSA-mobility and SALSA-work scores, as well as with the JTT on the dominant hand (**Figure 4B-D**). We also found significant correlations between TUG and SALSA along with SALSA-mobility scores (**Figure 4E**). Finally, we found positive and significant correlations between the dominant and nondominant JTT results with the SALSA scales, SALSA-work, and-dexterity (**Figure 4F-H**).

**Figure 5.**
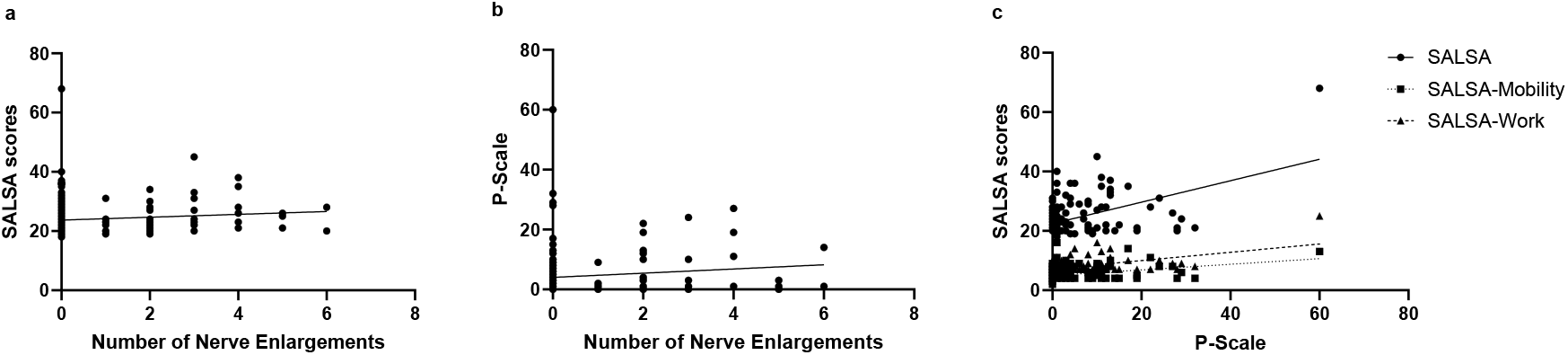
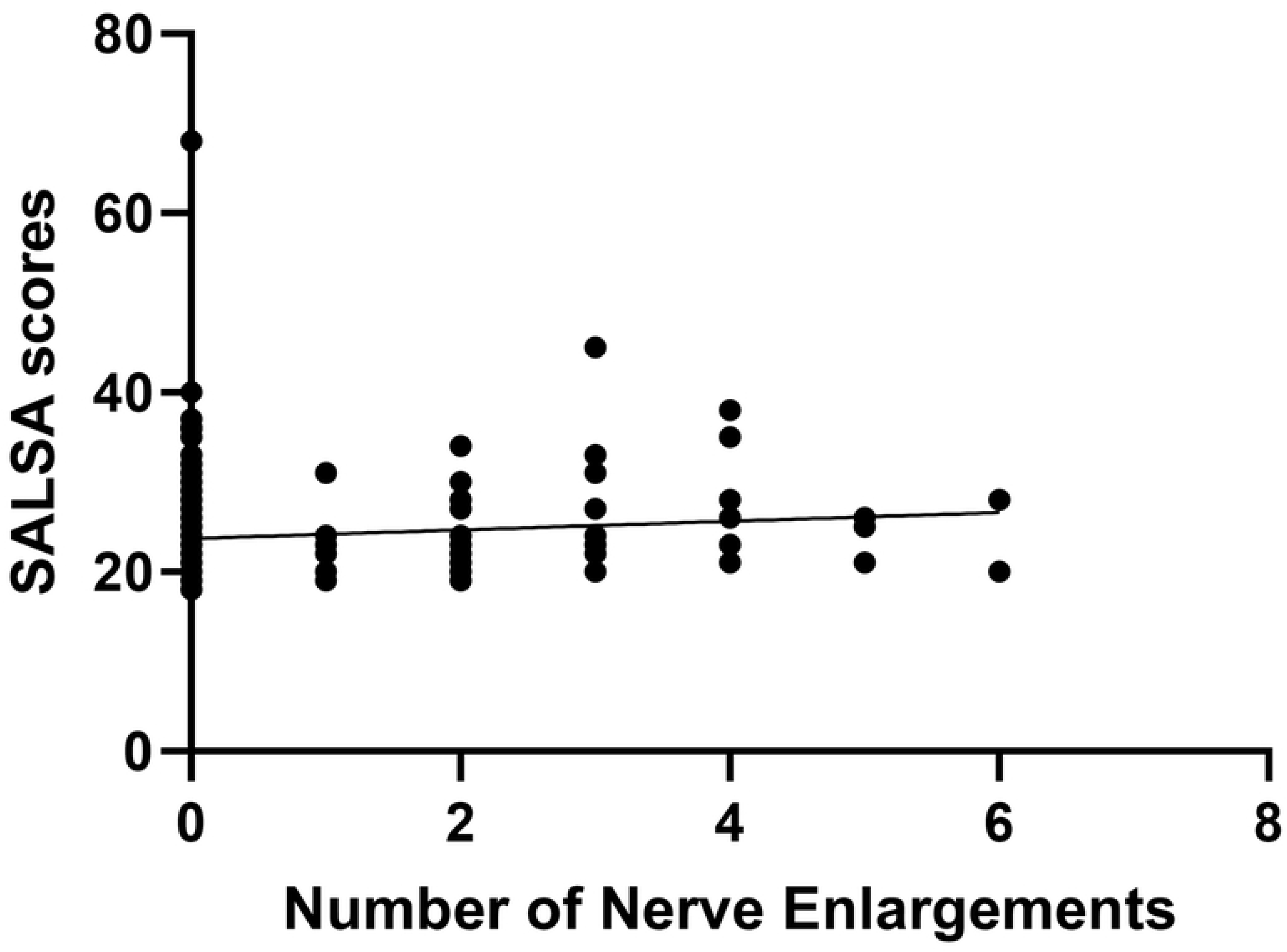
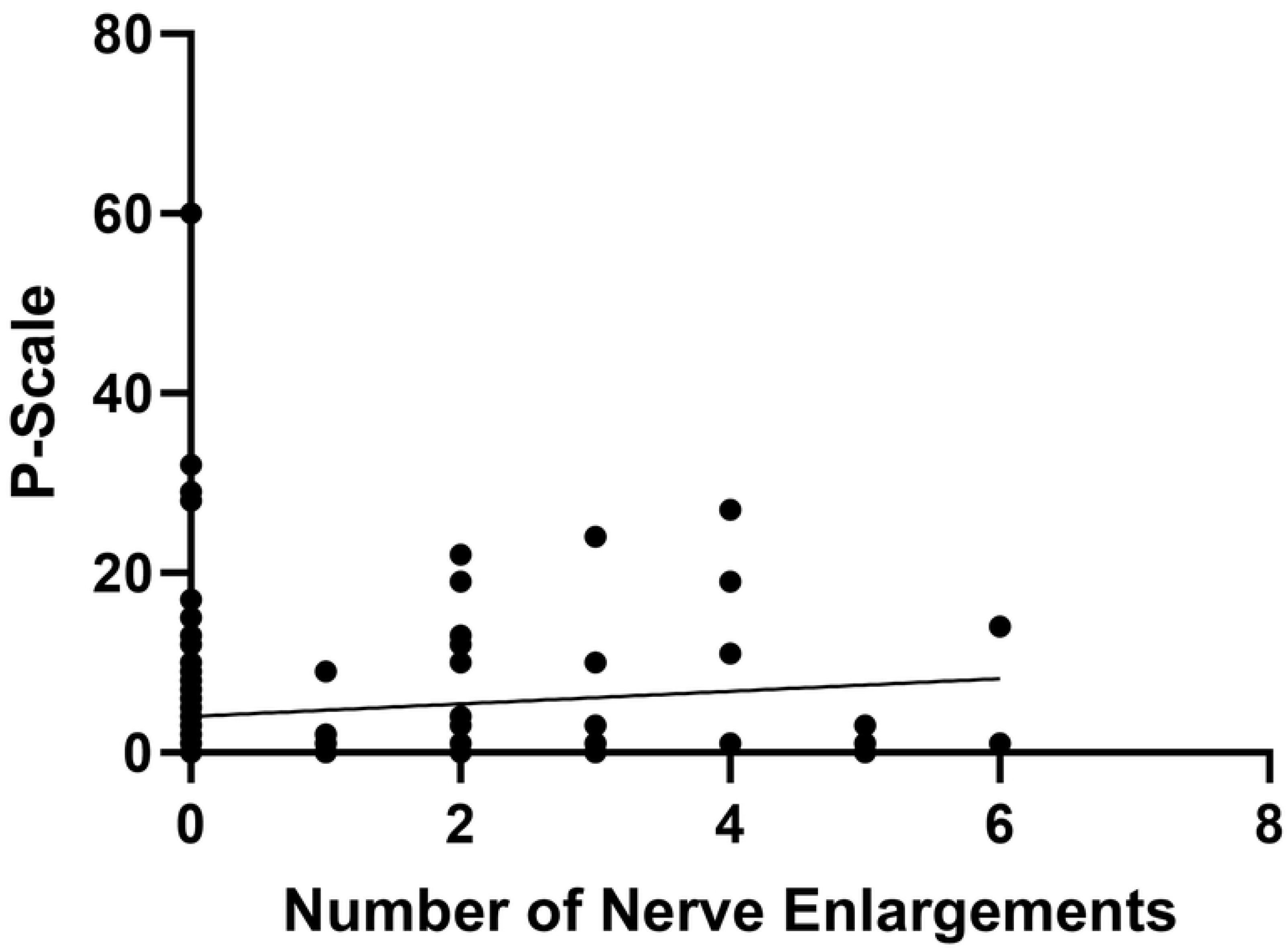
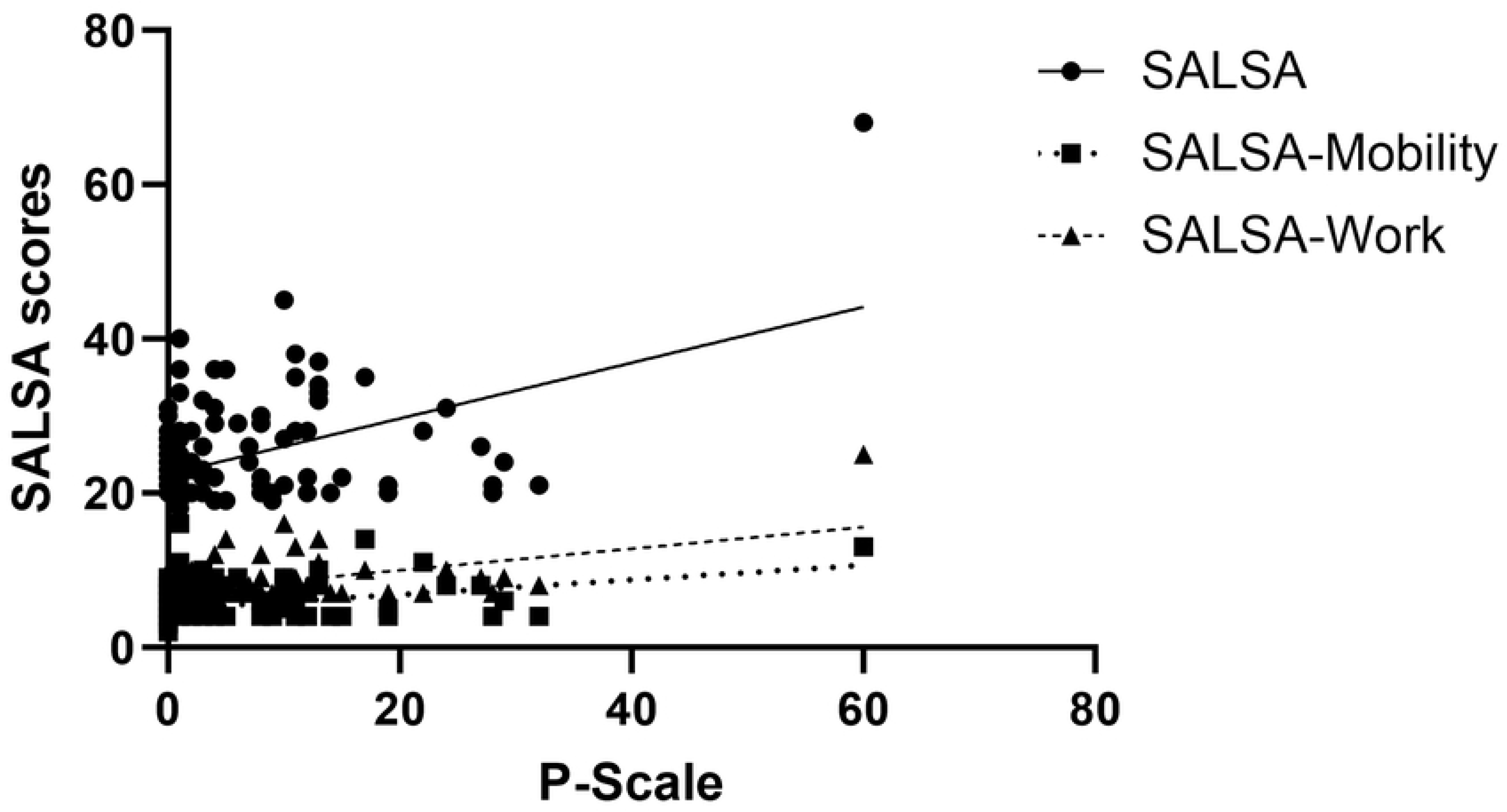
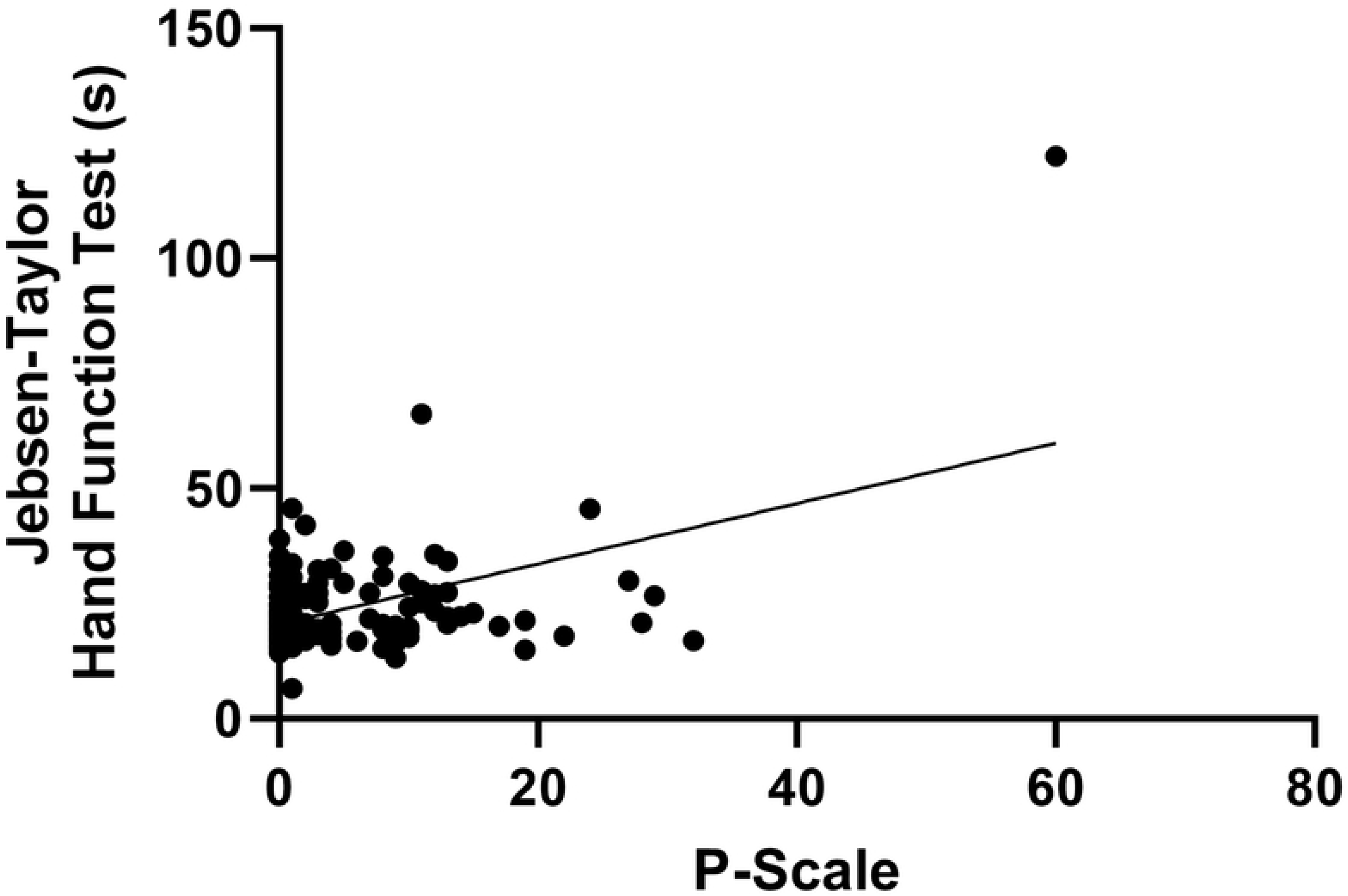
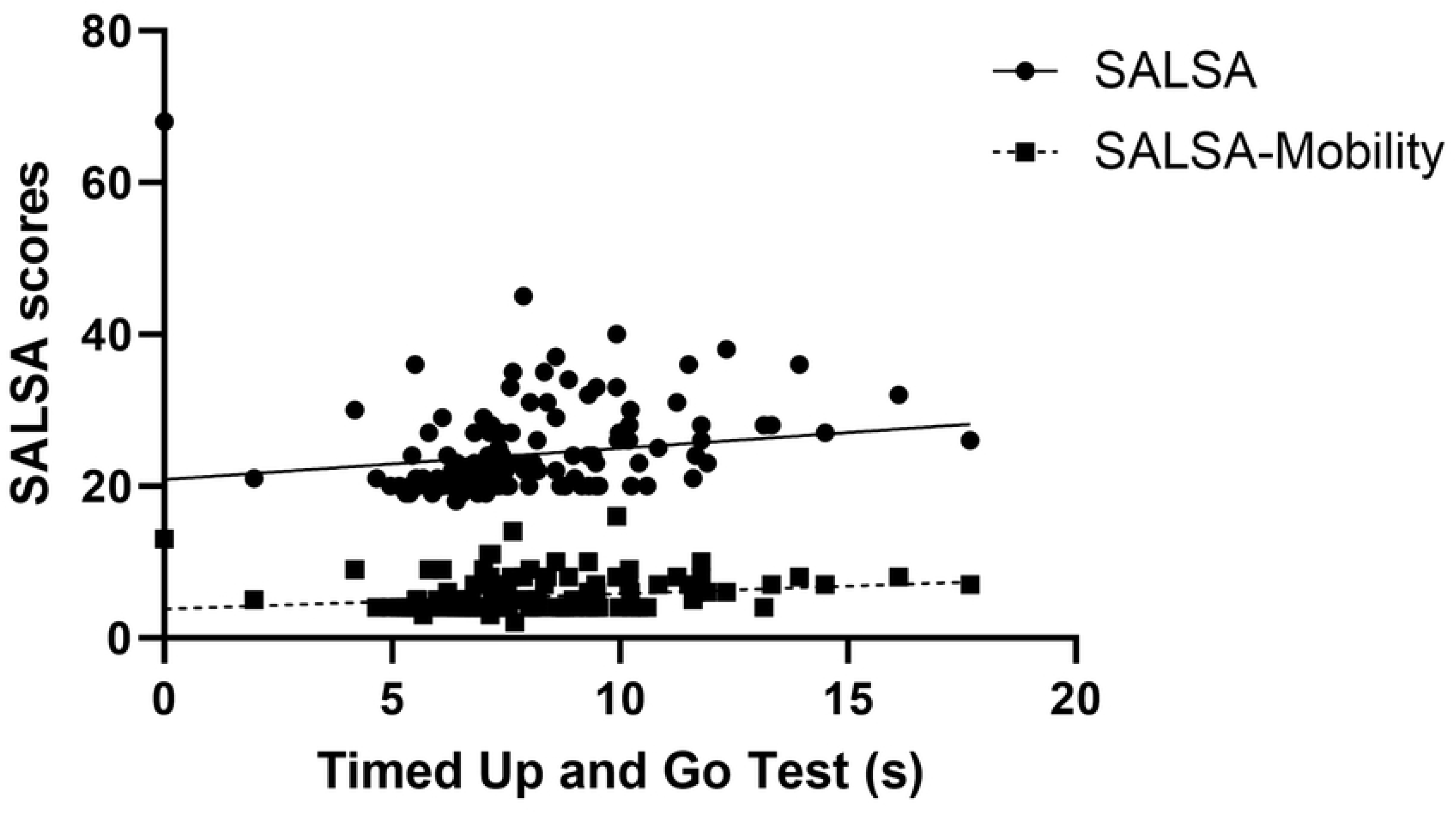
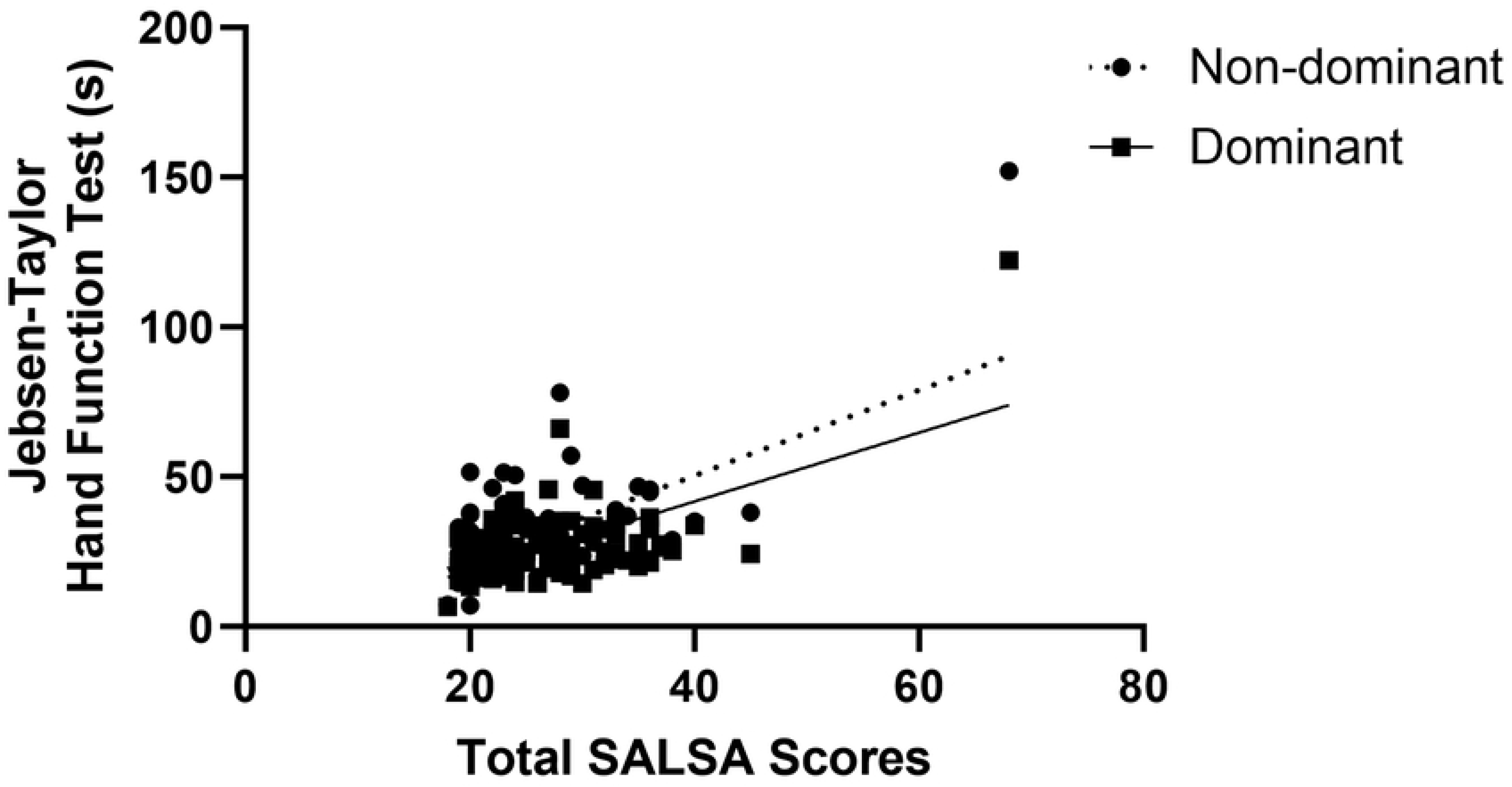
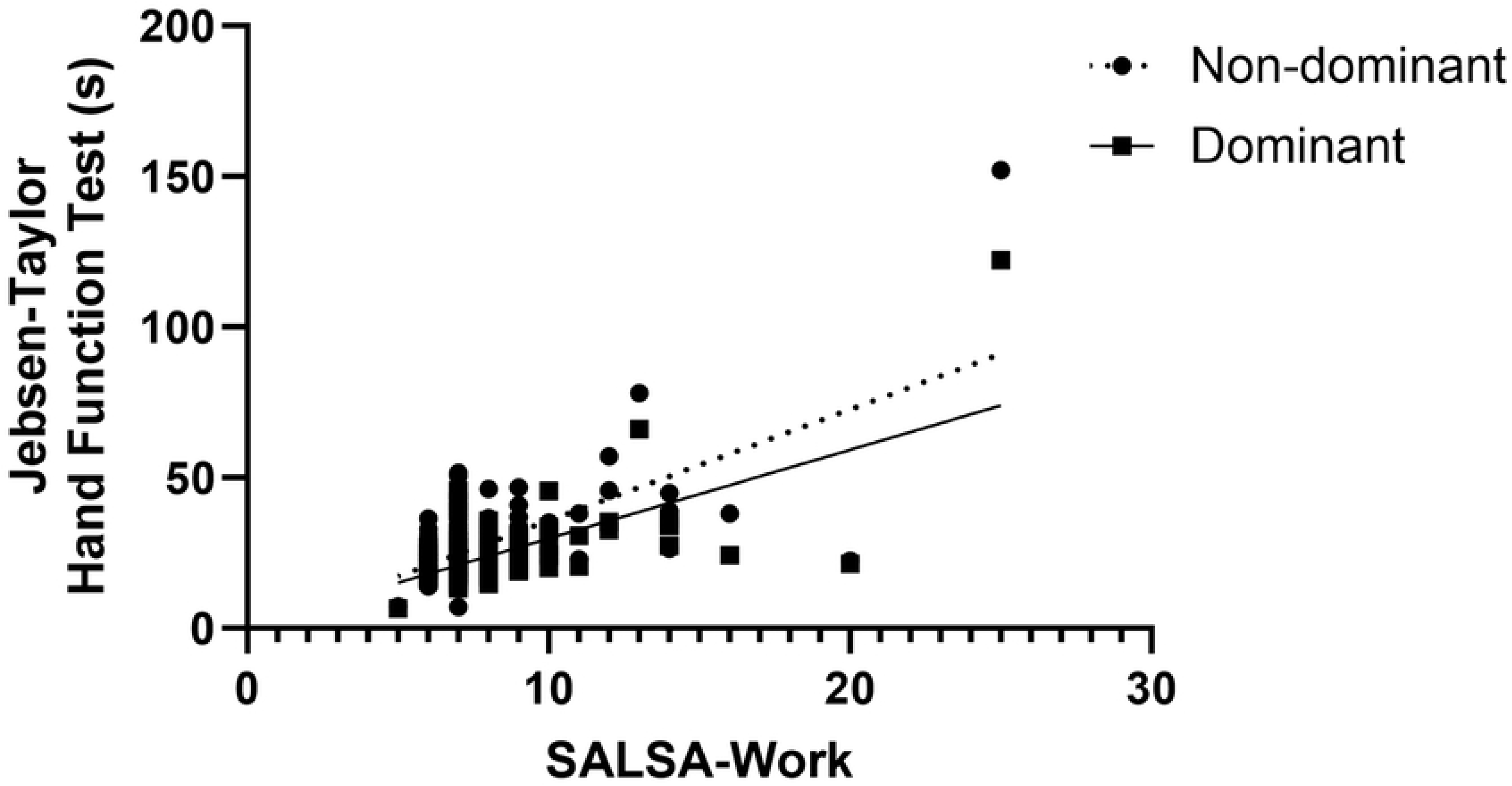
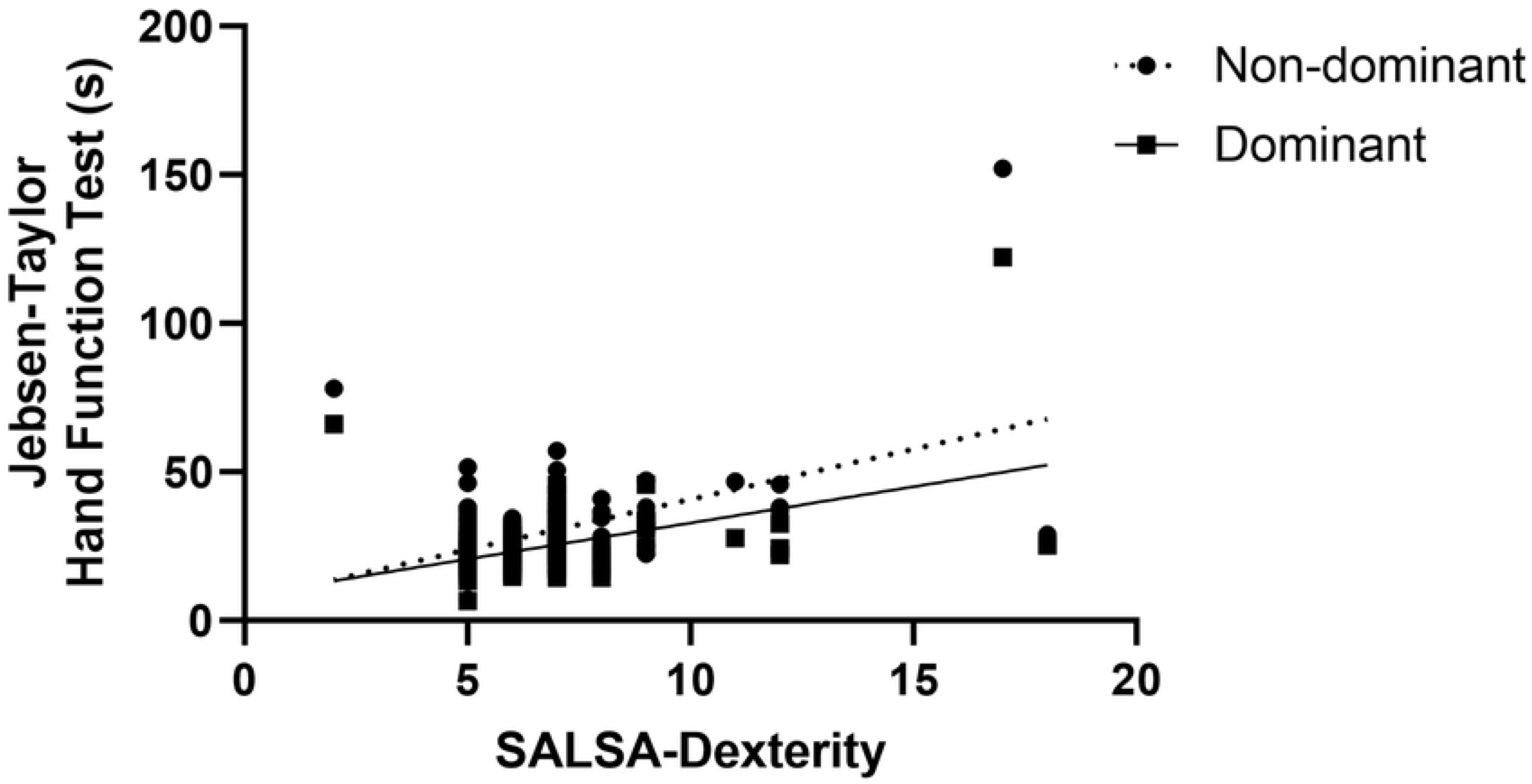
Correlations between the assessment tools according to the International Classification of Functioning, Disability and Health (ICF). (A) Health condition (number of nerve enlargement) with activity limitation (SALSA), Spearman’s r=0.1647, p=0.044. (B) Health condition (number of nerve enlargement) with participation restriction (P-scale), Spearman’s r=0.1673, p=0.0408. (C) Participation restriction (P-scale) and activity limitation (total SALSA (Spearman’s r=0.2722, p=0.0008), SALSA-mobility (Spearman’s r=0.2476, p=0.0023) and-work (Spearman’s r=0.3347, p<0.0001)). (D) Body function impairment (JTT in dominant hand) with participation restriction (P-scale), Spearman’s r=0.1742, p=0.033. (E) Body function impairment (TUG) and activity limitation in relation to lower extremities (total SALSA (Spearman’s r=0.4232, p<0.0001) and SALSA-mobility (r=0.3296, p<0.0001)). (F-H) Body function impairment (JTT) and activity limitation (total SALSA (Spearman’s r=04296, p<0.0001 and Spearman’s r=0.4659, p<0.0001), SALSA-work (Spearman’s r=0.3967, p<0.0001 and Spearman’s r =0.4236, p<0.0001), and SALSA-dexterity (Spearman’s r=0.4212, p<0.0001 and Spearman’s r=0.3737, p<0.0001)) related to upper extremities in non-dominant and dominant hands, respectively.

## Discussion

One of the purposes of this study is to utilize ICF to assess the functional disabilities experienced by individuals diagnosed with leprosy in Likupang, North Minahasa and Lewoleba, Lembata, Indonesia. ICF recognizes several dimensions of disability, including impairment in body structure and function, restrictions in activity, and limitations in participation. Additionally, the classification acknowledges the significant influence of physical and social environmental factors on the outcomes of disability. In accordance with this, our results imply potential relationships between each domain to one another in leprosy patients.[4]

Several personal factors for leprosy and leprosy-related disabilities have been studied. A meta-analysis of 17 studies found that leprosy was associated with male gender, performing manual labour, ever food-shortage, sharing household with another leprosy patient, and communal living with a minimum of 5 occupants.[30] Moreover, risk factors for leprosy-related disabilities include advanced age, male, low education, multibacillary leprosy type, the presence of leprosy reactions, nerve damage, delayed diagnosis, and treatment dropout.[31-36]

Findings related to demographic factors in this study, which seemed to affect the domains of ICF disability, have been observed in other studies. Most of the patients in this study were males within the range of productive age group, as also found in other studies from Indonesia, Brazil, and India.[37-40] Our results also showed that body function, activity limitation, and participation restriction had positive correlations with age. Higher male population in leprosy cases could be attributed to differences in sociocultural behaviours between genders and life choices that would lead to higher risk of contracting the infection.[41] More than half of this study population also had low educational attainment, indicating a shared context of socioeconomic vulnerability. This trend has been observed in Indonesian, Indian, and Brazilian studies by Menaldi et al. (2022), Karotia (2022), and Dergan et al. (2023), respectively. Unfortunately, diagnostic timepoints and treatment dropout were not explored.[8,40,42]

Leprosy patients with multi-bacillary type demonstrate more pronounced physical dysfunction and experience greater bodily pain when compared to the pauci-bacillary group, which primarily stems from nerve impairment in leprosy patients.[43] Multi-bacillary patients were significantly at higher risk of developing nerve-function impairment.[44] Additionally, a study by Lustosa et al. (2011) revealed that multibacillary patients were associated with lower SF-36 scores across four domains: physical functioning, bodily pain, general health perceptions, and social functioning. These patients were also at higher risk of being found with grade II disability at the time of diagnosis.[45]

Increased nerve thickening has been found to play a role in delaying diagnosis and potentially serves as a risk factor for deformity development.[46] Deformities were also more often seen in those with more than 3 thickened nerves. Our bivariate analyses found that the number of nerve enlargement in leprosy patients has weak but positive correlations to functional activity limitation and social participation restriction, which was represented by SALSA scale (r=0.165, p=0.044) and P-scale (r=0.167, p=0.041), respectively. It has been studied that nerve enlargement can lead to neuropathic pain due to entrapment syndrome. While it may not be the sole cause, nerve enlargement still contributes significantly to nerve pain occurrence.[47] A recent Brazilian study also suggested that leprosy patients experiencing nerve pain exhibited higher SALSA scores compared to those without pain.[48]

There were 29.33% of participants who experienced mild (27.33%), moderate (1.33%), and extreme (0.67%) activity limitations. This proportion is lower than that reported by Shivanna et al. (2022) in India, Nascimento et al. (2020) in Brazil, and Abdela et al. (2020) in Ethiopia. In two studies (Shivanna et al. and Nascimento et al.), most participants only felt mild functional limitation. High proportion of patients with activity limitations may indicate prior delayed diagnosis, inadequate management, and lack of physical rehabilitation services.[49,50,51]

Patients in this study who had limitations in social participation accounted for only one-eighth of the total participants, with 6.67% experiencing mild restriction, 4% experiencing moderate restriction, and 0.67% experiencing extreme restriction. These figures are notably lower than those found in studies conducted in Brazil, Ethiopia, and Nigeria, which reported participation restriction rates of 24%, 55.1%, and 89.3%, respectively.[50,51,52] Variation in cultural characteristics, socioeconomic conditions, stigma, and availability of rehabilitative services may account for these differences.[50]

Functional limitation and societal restriction are impacted by many factors. According to a previous study by Nogueira in Fortaleza, most of elderly people with leprosy can still do basic daily tasks such as opening bottles with screw caps, cooking, carrying heavy objects, and walking uneven ground.[53] Stigma and prejudice associated with leprosy can lead to societal exclusion even in the absence of visible lesions or impairments.[50] Several factors such as self-stigmatization, activity limitations, family-related issues, poverty, low education levels, inadequate rehabilitation services that includes community-based rehabilitation programs and community ignorance about the disease and its transmission can affect societal participation restriction.[51]

This study also found significant differences of SALSA scores according to the degree of WHO disability for hands and feet (p=0.0012 and p=0.0003, respectively). Similar findings from de Souza et al. (2016) indicated that SALSA scores are associated with the degree of impairment (p<0.01).[24]

There were significant differences between total participation scale and hand disability based on WHO disability grading (p=0.0087). Grade 2 disabilities, particularly those affecting the hands and especially with deformities and resulting amputations can cause discomfort and limit patients’ ability to interact with their environments, participate in community activities, and pursue employment opportunities. This may lead to reduced autonomy outside the home and interference with family roles.[54]

Bivariate analyses also found significant positive corelations between JTT in both dominant and non-dominant hands, and SALSA scores, including the work and dexterity domains, suggesting that reduced hand function is paralleled with limited functional activity. Meanwhile, TUG test was used to determine functional mobility, which is typically reduced in people with foot disability.[55] There were positive correlations between TUG test and both SALSA and mobility domain of SALSA scale. TUG test and total participation scale denoted no significant correlation with p=0.3956. There were also no significant differences of total participation scale between different WHO foot disability grades (p=0.0515).

We found an interesting trend where hand function affected participation restriction whereas foot did not. In an older study of 63 patients with hereditary motor and sensory neuropathy (HMSN) or Charcot-Marie-Tooth disease (CMT), upper limb disability, and not lower extremity, was considered to correlate independently with participation restriction.[56] The latter has been linked to mortality and morbidity with age. We observe a similar trend of association between the hand function and societal participation in a recent study by Akbarfahimi et al. (2021) among 84 stroke patients.[57] Upper limb function measured using Fugl-Meyer scale was found to correlate significantly with participation score (r = 0.315, p

=0.003). In an older study with 2291 healthy participants above 50 years of age, lower extremity strength and balance were not significantly associated with societal participation.[58] However, the odds of having self-reported limitation in social participation were higher in those with slower gait speed or less than 1 m/s (odds ratio [OR] = 3.1 99% confidence interval [CI] 1.5–6.2) compared to those above. Some studies have even shown supporting evidence of relationship between gait speed and longer life. However, none of the studies discussed the reason behind this trend. Future studies detailing analysis of upper and lower extremity functions and disabilities are needed.

Most of the population in Likupang work as fishermen, which may be the cause of higher participation rates compared to studies in other countries.[59,60] Since fisherman’s work is rather self-reliant, they are not really affected by the stigma around them. However, fishermen required a fully function hands to maintain their job. Most of them use handline fishing, using hand grip and strength to be used. In addition to fishing, people of Likupang also work in other industry as well, such as farming and construction worker.[61, 62] The main occupation in Lembata island is corn farming industry and most of the farmers spent around 60% of their working time on the field.[61,63] Working in this industry demand people to have high functioning hands, due to its labor characterization of this job.[64] Farmers and fishermen with leprosy will suffer greatly mainly because their incapability to work and use their tools properly. This study showed a significant correlation between hand disability and participation scale, which include occupational activities such as fishing and farming.

Based on the ICF model, environmental and personal factors are also important components of disability. Risk factors for leprosy in Indonesia include floor height, house ventilation area, house window usage habits, floor and wall types, environmental sanitation, history of household contact, residential density, humidity, and economic status.[65,66,67] Humidity poses a significantly increased risk, with an 8.415-fold greater likelihood, of leprosy occurrence within the community. Similarly, personal hygiene presents a 6.926-fold higher risk of leprosy occurrence in the community. Residential density in Indonesian society is associated with a substantially elevated risk, estimated to be 5.754 times greater, of experiencing leprosy.[67]

The populations in Lewoleba and Likupang were unequally distributed with several areas exhibiting higher density than others. There were also inadequate access to clean water, insufficient drainage and sanitation systems, and relatively high levels of humidity, all of which are risk factors for leprosy as well as posing as further challenges in managing leprosy.[68,69] One possible approach to preventing leprosy in remote and rural areas is by providing public education on maintaining the physical condition of houses in accordance with established standards. This includes ensuring the presence of ceilings, utilizing easily cleanable flooring materials, maintaining comfortable humidity levels, and limiting the occupancy of bedrooms to a maximum of two individuals.[70] Conversely, marital status and proximity to healthcare facilities do not significantly affect the likelihood of leprosy-related disability.[33]

## Limitations

This study has several limitations. Firstly, this study only included the patients who were willing to participate and excluded those who would potentially be more reclusive and isolated. Secondly, the population included in the study also came from two different areas, each with unique social and environmental characteristics. There may be some gaps in demographical factors, which we could not currently analyze, that would influence the outcome of the study. Finally, although we found that the personal factors identified in our study such as age, gender, and educational background significantly contribute to leprosy-related disability, other influential factors including personal hygiene, contact history, socio-economic status, health beliefs, and religion were not specifically examined in our research,[66,71-73] highlighting the need for further exploration and investigation.

## Conclusion

There are significant and positive correlations between the body structure and body function impairment, activity limitation and participation restriction in individuals with leprosy in Indonesia, based on the ICF concept in mapping the functional disabilities. Similar trends are also found in body function and activity limitation, as well as between the hand function and participation restriction. Activity limitation and participation is lower in patients with more severe hand disability. In contrast, foot function and disability do not seem to affect societal participation.

## Data Availability

All relevant data are within the manuscript and its supporting information files.

## Acknowledgment

This work is supported by the Directorate of Research and Community Engagement of Universitas Indonesia. We would like to thank the directors and staffs of St. Damian Hospital Lewoleba-Lembata-East Nusa Tenggara, Ministry of Health East Lewoleba-East Nusa Tenggara, Ministry of Health North Minahasa-North Sulawesi, and Faculty of Medicine, University of Sam Ratulangi, Manado City-North Sulawesi. We would like to acknowledge the KATAMATAKU team and the medical students of Universitas Indonesia.

## Notes

### Competing Interest Statement

The authors have declared no competing interest.

### Funding Statement

Yes

### Author Declarations

The study protocol was approved by the Ethics Committee of Faculty of Medicine, Universitas Indonesia under ethical clearance ND-454/UN2.F1/ETIK/PPM.00.02/2022 and was designed in consideration of the principles proposed by the Helsinki Declaration.

